# Machine learning algorithm to perform ASA Physical Status Classification

**DOI:** 10.1101/2021.10.05.21264585

**Authors:** Alexander Pozhitkov, Naini Seth, Trilokesh D. Kidambi, John Raytis, Srisairam Achuthan, Michael W. Lew

## Abstract

**Background:** The American Society of Anesthesiologists (ASA) Physical Status Classification System defines peri-operative patient scores as 1 (healthy) thru 6 (brain dead). The scoring is used by the anesthesiologists to classify surgical patients based on co-morbidities and various clinical characteristics. The classification is always done by an anesthesiologist prior operation. There is a variability in scoring stemming from individual experiences / biases of the scoring anesthesiologists, which impacts prediction of operating times, length of stay in the hospital, necessity of blood transfusion, etc. In addition, the score affects anesthesia coding and billing. It is critical to remove subjectivity from the process to achieve reproducible generalizable scoring.

**Methods:** A machine learning (ML) approach was used to associate assigned ASA scores with peri-operative patients’ clinical characteristics. More than ten ML algorithms were simultaneously trained, validated, and tested with retrospective records. The most accurate algorithm was chosen for a subsequent test on an independent dataset. DataRobot platform was used to run and select the ML algorithms. Manual scoring was also performed by one anesthesiologist. Intra-class correlation coefficient (ICC) was calculated to assess the consistency of scoring

**Results:** Records of 19,095 procedures corresponding to 12,064 patients with assigned ASA scores by 17 City of Hope anesthesiologists were used to train a number of ML algorithms (DataRobot platform). The most accurate algorithm was tested with independent records of 2325 procedures corresponding to 1999 patients. In addition, 86 patients from the same dataset were scored manually. The following ICC values were computed: COH anesthesiologists vs. ML – 0.427 (fair); manual vs. ML – 0.523 (fair-to-good); manual vs. COH anesthesiologists – 0.334 (poor).

**Conclusions:** We have shown the feasibility of using ML for assessing the ASA score. In principle, a group of experts (*i*.*e*. physicians, institutions, *etc*.) can train the ML algorithm such that individual experiences and biases would cancel each leaving the objective ASA score intact. As more data are being collected, a valid foundation for refinement to the ML will emerge.

## Background

The American Society of Anesthesiologists Physical Status Classification System (ASA score) is used by anesthesiologists/proceduralists to classify surgical patients based on co-morbidities and other clinical characteristics. The grades of ASA scores range from 1 (healthy) to 6 (brain-dead organ donor) and are routinely assigned by individual anesthesiologists/proceduralists prior to the operative procedure (Table 1). This practice can result in a patient potentially receiving a different ASA score when assessed by a different anesthesiologists/proceduralists; and can result in downstream impact as a predictor of operating times, hospital length of stay, postoperative infection rates, necessity of blood transfusion and overall morbidity and mortality rates (1-5). In addition, the ASA score is used in the determination of anesthesia coding and billing. Because of the utility and the impact of the ASA score, it would be beneficial to devise a standardized method for its calculation.

**Table 1.** ASA scores, adopted from the American Society of Anesthesiologists

| ASA PS Classification | Definition | Adult Examples, Including, but not Limited to: | Pediatric Examples, Including but not Limited to: | Obstetric Examples, Including but not Limited to: |
| --- | --- | --- | --- | --- |
| ASA I | A normal healthy patient | Healthy, non-smoking, no or minimal alcohol use | Healthy (no acute or chronic disease), normal BMI percentile for age |  |
| ASA II | A patient with mild systemic disease | Mild diseases only without substantive functional limitations. Current smoker, social alcohol drinker, pregnancy, obesity (30<BMI<40), well-controlled DM/HTN, mild lung disease | Asymptomatic congenital cardiac disease, well controlled dysrhythmias, asthma without exacerbation, well controlled epilepsy, non-insulin dependent diabetes mellitus, abnormal BMI percentile for age, mild/moderate OSA, oncologic state in remission, autism with mild limitations | Normal pregnancy*, well controlled gestational HTN, controlled preeclampsia without severe features, diet-controlled gestational DM. |
| ASA III | A patient with severe systemic disease | Substantive functional limitations; One or more moderate to severe diseases. Poorly controlled DM or HTN, COPD, morbid obesity (BMI ≥40), active hepatitis, alcohol dependence or abuse, implanted pacemaker, moderate reduction of ejection fraction, ESRD undergoing regularly scheduled dialysis, history (>3 months) of MI, CVA, TIA, or CAD/stents. | Uncorrected stable congenital cardiac abnormality, asthma with exacerbation, poorly controlled epilepsy, insulin dependent diabetes mellitus, morbid obesity, malnutrition, severe OSA, oncologic state, renal failure, muscular dystrophy, cystic fibrosis, history of organ transplantation, brain/spinal cord malformation, symptomatic hydrocephalus, premature infant PCA <60 weeks, autism with severe limitations, metabolic disease, difficult airway, long term parenteral nutrition. Full term infants <6 weeks of age. | Preeclampsia with severe features, gestational DM with complications or high insulin requirements, a thrombophilic disease requiring anticoagulation. |
| ASA IV | A patient with severe systemic disease that is a constant threat to life | Recent (<3 months) MI, CVA, TIA or CAD/stents, ongoing cardiac ischemia or severe valve dysfunction, severe reduction of ejection fraction, shock, sepsis, DIC, ARD or ESRD not undergoing regularly scheduled dialysis | Symptomatic congenital cardiac abnormality, congestive heart failure, active sequelae of prematurity, acute hypoxic-ischemic encephalopathy, shock, sepsis, disseminated intravascular coagulation, automatic implantable cardioverter-defibrillator, ventilator dependence, endocrinopathy, severe trauma, severe respiratory distress, advanced oncologic state. | Preeclampsia with severe features complicated by HELLP or other adverse event, peripartum cardiomyopathy with EF <40, uncorrected/decompensated heart disease, acquired or congenital. |
| ASA V | A moribund patient who is not expected to survive without the operation | Ruptured abdominal/thoracic aneurysm, massive trauma, intracranial bleed with mass effect, ischemic bowel in the face of significant cardiac pathology or multiple organ/system dysfunction | Massive trauma, intracranial hemorrhage with mass effect, patient requiring ECMO, respiratory failure or arrest, malignant hypertension, decompensated congestive heart failure, hepatic encephalopathy, ischemic bowel or multiple organ/system dysfunction. | Uterine rupture. |
| ASA VI | A declared brain-dead patient whose organs are being removed for donor purposes |  |  |  |

Machine learning algorithms in artificial intelligence are designed to identify patterns in complex datasets, such as clinical patient data (6, 7). One proposal to improve the ASA scoring algorithm is the implementation of machine learning techniques to refine and automate the computation of the ASA score. From applying machine learning in clinical data collection, multiple patient characteristics can be distilled into an objective and consistent ASA score. Furthermore, the analysis will be able to extract the relationship between the co-morbidities and the assigned ASA scores, and therefore can develop a standard method of assigning ASA scores prospectively.

According to the American Cancer Society in the United States, cancer is the second leading cause of death in men and women 45-64 years of age and the prevalence is projected to increase from 16.9 million in 2019 to 22.1 million cancer survivors in 2020 (10). Considering that oncology is a specialized niche, the familiarity with the type of cancer, its stage, the therapeutic regimens in conjunction with the presence of a coexisting disease(s), the opportunity persists to encounter even a higher variability in the ASA score.

To that point, we developed a machine learning algorithm that predicts ASA scores with a fair degree of accuracy for the cancer patient.

## Methods

City of Hope (COH) is a comprehensive cancer center and all patients in our database has cancer or is a cancer survivor. The ASA score from the anesthesiologist to the assigned surgical procedure (19,095 procedures corresponding to 12,064 patients (some patients had more than one surgery)) from December 2, 2017 to April 30, 2020 was collected. The clinical characteristics, laboratory values and physician notes were extracted from the City of Hope Enterprise Data Warehouse (COH-EDW), which is a MS SQL database mirroring the data from Epic electronic medical records system. A custom-made SQL query was created to extract relevant attributes. A DataRobot machine learning platform was used to automatically select, train and validate a machine learning algorithm associating ASA score with clinical characteristics. The analysis was programmed as a multiclass classification problem. The ASA scores were compared to each other. In a subset of 86 random patients a single anesthesiologist (M. Lew) was blinded to the assigned ASA score determined at the time of surgery; and assigned an ASA score based solely on coded diagnosis(s). M. Lew’s ASA score was compared to: 1) the assigned ASA score at the time of surgery and 2) the ASA score determined by the machine learning algorithm.

## Results and Discussion

The following attributes were extracted: patient’s age at the time of surgery, gender, STOP BANG score (8), stress test results (specifically if ischemia or reversible ischemia occurred), medication within 30 days prior surgery, abnormal laboratory values within 30 days of surgery, and ICD10 codes for diagnoses.

The following pharmacological classes were considered: opioids, antacids, antianginal agents, antiarrhythmic, antiasthmatic, anticoagulants, anticonvulsant, antidiabetic, antihypertensive (i.e beta blockers, calcium blockers), antimyasthenic agents, antiparkinsonian, antipsychotics, diuretics, endocrine (i.e. thyroid replacement, corticosteroids), vasopressors.

The following laboratory values were considered as relevant: were albumin, alanine transaminase, aspartate aminotransferase, bicarbonate, bilirubin, brain natriuretic peptide, calcium (serum and urine),, creatine kinase, creatinine, glucose, hematocrit, hemoglobin, INR, prothrombin time, potassium (serum and urine), sodium (serum and urine), platelets, partial thromboplastin time, T3, T4, troponin, thyroid-stimulating hormone, Von Willebrand factor, white blood cell count. A laboratory value was considered abnormal low/high if within 30 days prior surgery it was measured below or/and above the reference values, respectively.

The above-described variables are a significant extension to the list of variables used in a recent ASA estimation work done by Zhang et al. (7).

### Training

A machine learning algorithm was trained with a dataset comprising 19,095 records corresponding to 12,064 patients (some patients had more than one surgery) from December 2, 2017 to April 30, 2020 (Supplementary file dataset20210302_clean.tsv). The DataRobot platform automatically allocated a subset of the training dataset for validation and holdout. Several machine learning algorithms were set to compete, and one final algorithm was selected as the most accurate (Figure 1). Upon completion of the training, for the holdout AUC = 0.76. Relative impact of the attributes is shown on Figure 2.

**Figure 1.**
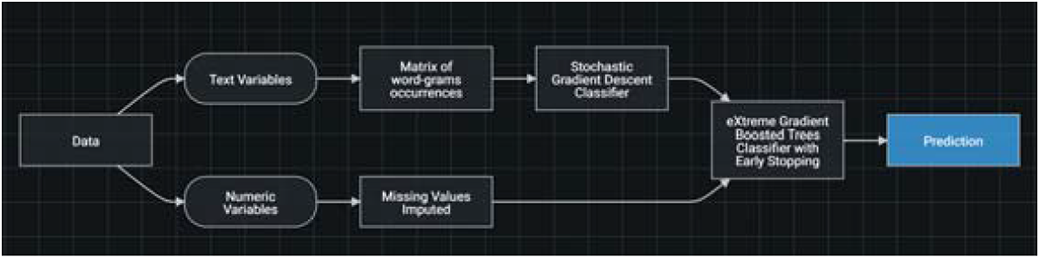
Schematic diagram of data processing for the machine learning algorithm.

**Figure 2.**
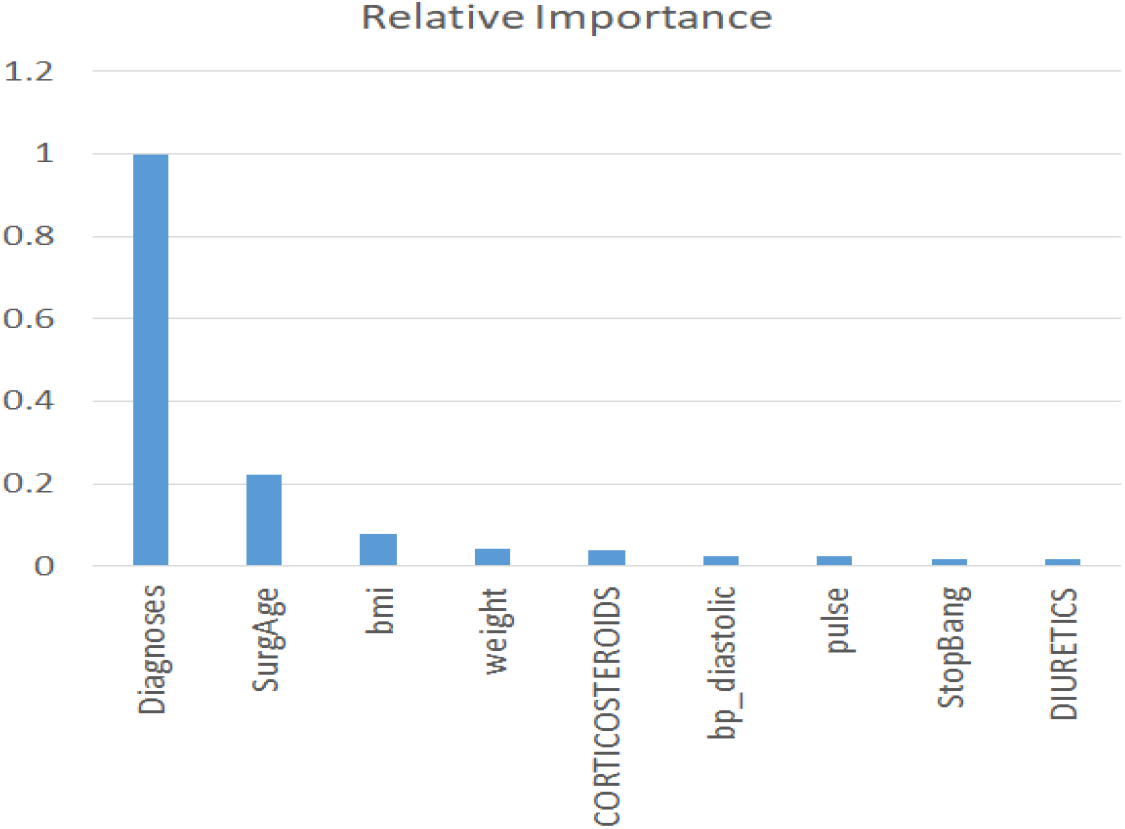
Relative importance of the patient attributes for the prediction algorithm.

### Prediction

The 3-months period of Jan 5, 2020 to May 8, 2020 was used to perform a validation experiment; the number of records was 2325 corresponding to 1999 patients (Supplementary file asa_may1Aug5_clean.tsv). Predictions were made by the trained model (above) in the form of weights assigning a case to one of the ASA score, 1-6. A class with maximum weight was considered as the predicted ASA score (see Supplementary file coh_vs_dr_full.xlsx). To assess the consistency of predictions, an intraclass correlation coefficient (ICC) was computed (9) to be 0.427 (0.393, 0.459). This ICC value corresponds to “fair” consistency between ASA scoring performed by the machine-learning algorithm and the COH anesthesiologists.

We took a random subsample of 86 patients and scored them manually (by M. Lew) based on the same attributes that were provided to the DataRobot (see Supplementary file coh_ml_dr_subset.xlsx). The ICC between M. Lew and DataRobot scoring is 0.523 (0.351, 0.66), which can be considered “fair to good” and apparently higher than the ICC reported above. Interestingly, the consistency between M. Lew and COH anesthesiologists scoring was found “poor”: ICC 0.334 (0.133, 0.509).

### Interpretation

The ASA scoring system inevitably introduces subjectivity when it is done by an anesthesiologist/proceduralist, because there is an element of human interaction between the physician and the patient during the time of scoring. Not only the medical history and answers to pre-defined questions are considered, but also actual symptoms, feedback, and overall disposition of the patient. At the same time, physicians experience “fatigue” as they score numerous patients in their day-to-day practice. Given that the COH dataset is composed from the scorings of as many as 17 anesthesiologists, it is reasonable to believe that the subjectivity was effectively nullified. Hence, the machine was trained on perhaps not entirely objective, but certainly consensus scores. Furthermore, one refinement during the process was that scores predicted by the algorithm which varied from the clinician was reviewed by one anesthesiologist (decreasing practitioner variability) to further improve the consistency and the accuracy of the machine learning model.

It is interesting to note that the machine scorings for the data of the 3-months period were in fair-to-good agreement with both the COH anesthesiologists and M. Lew’s scorings; however, M. Lew’s scoring was further away from COH anesthesiologists as a group. This can be explained by the fact that M. Lew only reviewed the coded diagnosis(s) and lacked the ability to have the personal interaction in real time with the patients. Our findings indicate that the machine was not far off from both the consensus (COH anesthesiologists) and individual physician (M. Lew) scoring.

## Conclusions

The ASA scoring process is not a simple set of rules that one could codify and follow; especially as it relates to the oncology patient. As a comprehensive cancer center, we feel confident that the machine learning algorithm addresses more granular data compared to scoring cancer as one diagnosis. Ultimately a group of “experts” (*i*.*e*. physicians, institutions, healthcare networks, and/or payors) can develop qualifying data, train the machine, and formulate an objective ASA score. As data collection increases, it will provide a valid foundation for refinement to the machine learning ASA scoring algorithm. This is the first known study to apply machine learning to the cancer patient with the goal of eliminating the subjectivity of the ASA score as well as improving the granularity.

## Supporting information

Supplementary file coh_ml_dr_subset.xlsx

Supplementary file coh_vs_dr_full.xlsx

## Data Availability

Raw and processed data are available as supplementary files.

## Declarations

### Ethics approval and consent to participate

The study was approved by the COH institutional review board under IRB #17467 “Correlation of Patient Characteristics and Assigned American Society of Anesthesiologist Risk Assessment Scores Using a Machine Learning Algorithm”.

### Consent for publication

Not applicable.

### Availability of data and materials

The raw data are available as supplementary files.

### Authors’ contributions

AP, MWL performed analysis and wrote the manuscript; NS, JR collected the data and researched records; TDK, SA contributed with concepts.

## List of Abbreviations

ML: machine learning
EDW: enterprise data warehouse
COH: City of Hope
ICC: intraclass correlation coefficient
ASA: American Society of Anesthesiologists

